# Persistent Homology of Tumor CT Scans is Associated with Survival In Lung Cancer

**DOI:** 10.1101/2020.12.06.20244863

**Authors:** Eashwar Somasundaram, Adam Litzler, Raoul R. Wadhwa, Steph Owen, Jacob G. Scott

**Affiliations:** Case Western Reserve University School of Medicine, Cleveland, OH 44195, USA; University of Colorado Boulder, Department of Applied Mathematics, Boulder, CO 80309, USA; Cleveland Clinic Lerner College of Medicine, Case Western Reserve University, Cleveland, OH 44195, USA; Lerner Research Institute, Department of Translational Hematology and Oncology Research, Cleveland, OH 44195, USA; Taussig Cancer Institute, Department of Radiation Oncology, Cleveland, OH 44195, USA

## Abstract

**Purpose:** Radiomics, the objective study of non-visual features in clinical imaging, has been useful in informing decisions in clinical oncology. However, radiomics currently lacks the ability to characterize the overall topological structure of the data. This niche can be filled by persistent homology, a form of topological data analysis that analyzes high-level structure. We hypothesized that persistent homology could be applied to lung tumor scans and analyze clinical outcomes.

**Methods:** We obtained segmented computed tomography lung scans (n × 565) from the NSCLC-Radiomics and NSCLC-Radiogenomics datasets in The Cancer Imaging Archive. For each scan, a cubical complex filtration based on Hounsfield units was generated. We calculated a feature curve that plotted the number of 0 dimensional topological features against each Hounsfield unit. This curve’s first moment of distribution was utilized as a summary statistic to predict survival in a Cox proportional hazards model.

**Results:** After controlling for tumor image size, age, and stage, the first moment of the 0D topological feature curve was associated with poorer survival (HR × 1.118; 95% CI × 1.026-1.218; p × 0.01). The patients in our study with the lowest first moment scores had significantly better survival (1238 days; 95% CI × 936-1599) compared to the patients with the highest first moment scores (429 days; 95% CI × 326-601; p × .0015).

**Conclusions:** We have shown that persistent homology can generate useful clinical correlates from tumor CT scans. Our 0-dimensional topological feature curve statistic predicts survival in lung cancer patients. This novel statistic may be used in tandem with standard radiomics variables to better inform clinical oncology decisions.

## 1 Introduction

The use of radiomics in tumor imaging has been an increasingly popular area of research to better inform cancer diagnosis, treatment, and prognosis^1^. Geometric properties of tumors such as size, surface area, and volume are commonly studied features among more non-visual features such as texture^2^. We propose extending the utility of radiomics by studying topological properties. In contrast to the local structural focus of geometry, topology focuses more on global structure.

Since topological properties would focus on general tumor shape properties, they are theorized to be more robust to noise and could capture information not found in traditional radiomic features. Intuitively, one can describe several topological differences between malignant and benign tumors. Benign tumors are well-connected and have a homogeneous shape whereas malignant tumors are more likely to have diffuse spread and necrotic cavities^3^. Quantifying the number of topological features may be useful in predicting patient survival. In fact, pathology already uses shape properties in the context of Gleason scores, which is a measure of prostate cancer severity based on prostate gland shape^4^.

Persistent homology is a popular technique in the -omics sciences to describe the topological features of large data sets. Persistent homology has already been used in cancer biology from genomics to histology^5,6^. A statistical measure inspired by persistent homology called the smooth Euler characteristic has been developed to predict clinical outcomes from glioblastoma tumor imaging^7^. Persistence images, an alternative representation of persistent homology, have been used in machine learning models to classify MRI images of hepatic tumors^8^. While radiomics and persistent homology have largely existed in separate worlds, they share similar challenges. In radiomics, one challenge is to find the most informative image features for analysis^9^.

In persistent homology, a similar challenge is finding the most useful topological data representation for visualization, statistical comparison, and predictive modeling. We were interested in whether image features related to topology in lung CT scans were associated with survival. Since we hypothesized earlier that an increased number of topological features may correlate to a more malignant tumor, we wanted to create a topological data representation that captured the quantity of topological features of a CT tumor scan. In this project, we create a new radiomics variable using the statistical summary variables of the 0D topological feature curve, our method of representing topological feature quantity. We describe this further in section 2. To our knowledge, no work has been done in using persistent homology to characterize survival in cancer patients using such a summary statistic. We show through a discrete and continuous analysis that moment 1 of our 0D topological feature curve (i.e. average number of 0D features) is significantly associated with worse survival.

## 2 Persistent Homology Background

Topological data analysis (TDA) encompasses a broad set of techniques used to highlight topological patterns in data. Computing persistent homology using cubical complexes is a popular method within TDA for describing topology in imaging data^10^. Counting topological features on an image requires a binary black and white image, so we cannot count the topological features from the grayscale CT scan directly. However, we can convert the CT scan into a series of binary black and white images (i.e. filtration) from which the topological features can be counted. In the context of persistent homology, cubical complexes are essentially synonymous with these binary images.

For each CT scan, we create a series of binary images (i.e. cubical complexes), one for each Hounsfield unit filtration value. For example, if the filtration value is -900 Hounsfield units, then all pixels with units less than or equal to -900 are colored black, and all pixels above are colored white. Topological features can be counted from these binary images. A particular topological feature can be described by dimension and the range of Hounsfield unit filtration values across which the feature is found.

We show this filtration process with slice 13 of scan 1 in **Figure 1A**. This slice is shown as it demonstrates a potential connection between tissue characteristics (calcifications) and topological features. At low Hounsfield unit thresholds, the image is mostly white. Islands of black pixels are considered connected components or 0D features. Tissue necrosis may result in a more fragmented tumor, which would increase the number of 0D features. “Lakes” of white pixels are considered holes or 1D features. Two-dimensional features are not shown in **Figure 1** as they only appear when considering the cubical complexes of all the slices stacked together as a three-dimensional structure. Intuitively, they can be thought of as volumetrically contiguous groups of white pixels existing in interior of the tumor image surrounded by black pixels. Calcifications would likely manifest as 1D or 2D features late in the filtration, which is shown when the Hounsfield unit threshold is 200 in **Figure 1A**.

**Figure 1.**
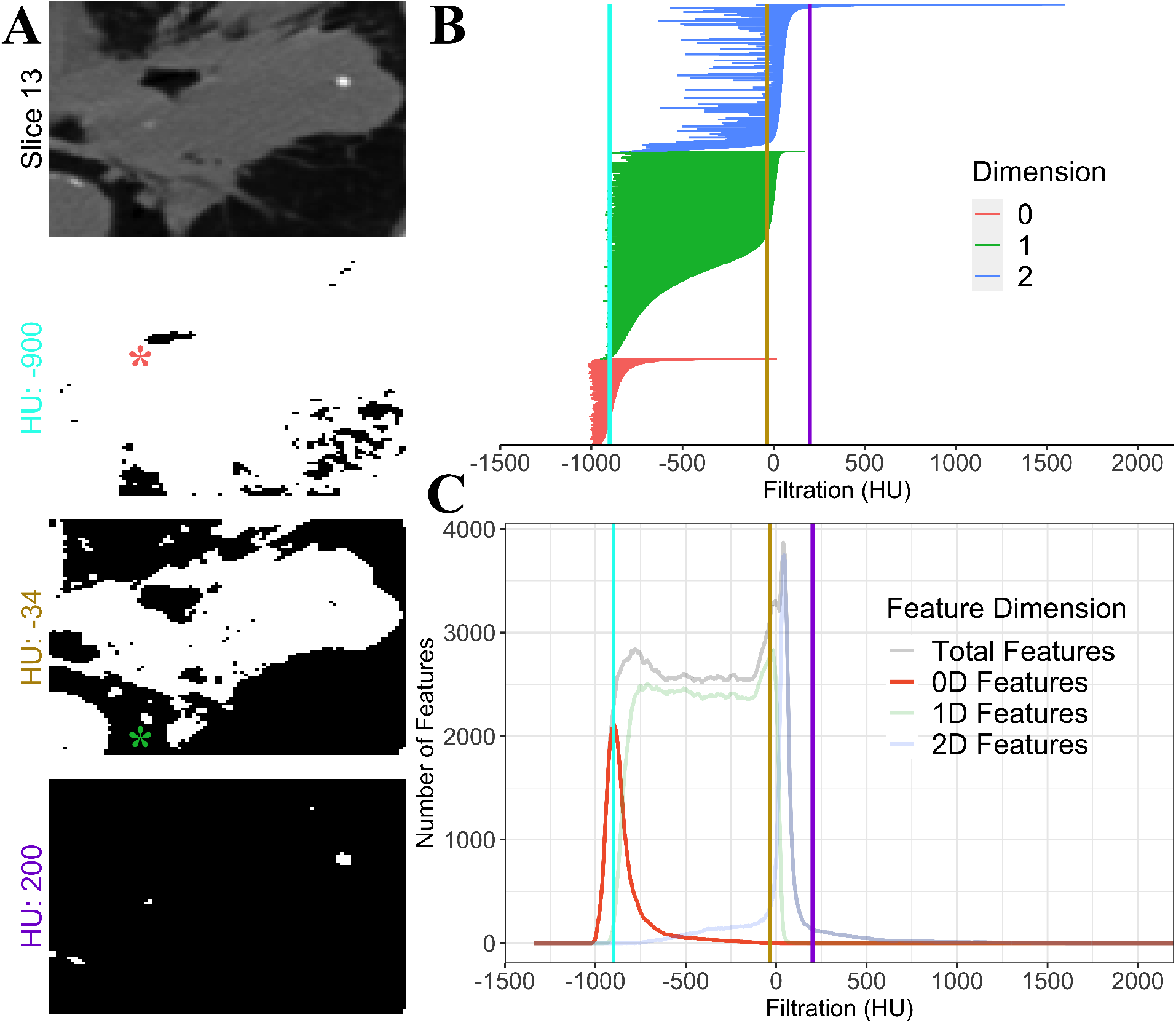
Methodology to generate cubical complexes. **(A)** An example slice from the Radiomics dataset. Each pixel value in a CT scan is described by a Hounsfield unit, which typically range from − 1024 to 3071. A cubical complex is created by selecting a Hounsfield value as a filter. Any pixel below or equal to this filter is colored black, and any pixel above is colored white, creating a binary image which becomes the cubical complex. Topological features can then be counted from each binary image. An example of a 0D feature is indicated by the red asterisk. An example of a 1D feature is indicated by the green asterisk. Two-dimensional features are not shown as they only appear when considering the cubical complexes of all the slices together as a three-dimensional structure. **(B)** We plot a barcode diagram representing the persistent homology of each topological feature. Color represents dimension, and the range represents the Hounsfield filtration unit range during which the feature existed. The colored vertical lines represent the Hounsfield units from the filtration values shown in the binary images. **(C)** We create three topological feature curves by summing the number of topological features by dimension at each Hounsfield unit filtration value. A fourth topological feature curve is generated by summing the total number of topological features regardless of dimension. The 0D topological feature curve is highlighted as we specifically use the moments of distribution of this curve for our survival analysis.

Topological barcodes are one of the most popular ways to visualize persistent homology^11^. Each topological feature in a barcode diagram is given a color to represent its dimension and a horizontal bar that spans the Hounsfield unit filtration values where the feature can be found. The barcode diagram in **Figure 1B** represents the entire cubical complex persistent homology of scan 1. Since we were interested in topological feature quantity, we developed an alternative representation of persistent homology called the topological feature curve.

The topological feature curve is a transformation of the barcode diagram that counts the number of bars (i.e. topological features) at each Hounsfield filtration value. The number of bars is then plotted against each Hounsfield unit filtration value. We can construct four topological feature curves, one for each feature dimension, plus a fourth curve that counts all features regardless of dimension. The topological feature curves in **Figure 1C** represent all feature curves in scan 1.

We used the raw moments of the distribution of the resulting 0D topological feature curve as predictor variables for our survival analysis. The exact mathematical calculation is described in **Supplemental Table 1**. We focused on the 0D curve’s characteristics because it showed the most variability across tumor scans. Incorporating all of the curves moments (16 potential variables) would not have been feasible given our sample size.

Similar topological summary statistics have been described in analyzing brain connectomes of individuals with Attention-deficit/hyperactivity disorder using 0D Vietoris-Rips complex persistent homology features^12^. Likewise, we also focus our analysis on 0D features; however, we generate our topological features using cubical complexes.

## 3 Results

**Table 1** gives a descriptive overview of the two lung scan data sets. These data sets were obtained from The Cancer Imaging Archive^13^. The NSCLC-Radiomics set (n × 421) had more patients compared to the NSCLC Radiogenomics set (n × 138)^14,15^.

**Table 1.** Descriptive statistics of the two cohorts of patient scans used in this study. All patient scans were downloaded from The Cancer Imaging Archive^13^. The moments of the 0D feature curves are presented as median [IQR] since the data was not normally distributed. Tumor image size was also not normally distributed and also presented as median [IQR]. All other data are presented as mean (SD) or as a proportion. Statistical comparisons between the two cohorts were performed using Wilcoxon rank-sum test for the non normally distributed data. Student’s t-test was used for the other continuous data. Chi-squared test was used for the categorical stage and sex data.

|  | Whole Cohort | NSCLC-Radiogenomics | NSCLC-Radiomics | p-value | Missing (%) |
| --- | --- | --- | --- | --- | --- |
| Total Sample Size | 559 | 138 | 421 |  |  |
| Vital Status (% Dead) | 424 (75.8) | 51 (37.0) | 373 (88.6) | <0.001 | 0.0 |
| Moment 1 | 24.23 [5.57, 70.40] | 21.07 [5.22, 69.19] | 26.26 [5.64, 71.48] | 0.782 | 0.0 |
| Moment 2 | $9.49 \times 10^3$<br>[$7.72 \times 10^2$,<br>$5.99 \times 10^4$] | $1.37 \times 10^4$<br>[$8.69 \times 10^2$,<br>$1.64 \times 10^5$] | $8.74 \times 10^3$<br>[$7.06 \times 10^2$,<br>$4.55 \times 10^4$] | 0.032 | 0.0 |
| Moment 3 | $3.92 \times 10^6$<br>[$1.31 \times 10^5$,<br>$6.56 \times 10^7$] | $9.12 \times 10^6$<br>[$1.63 \times 10^5$,<br>$4.21 \times 10^8$] | $3.29 \times 10^6$<br>[$9.85 \times 10^4$,<br>$4.16 \times 10^7$] | 0.003 | 0.0 |
| Moment 4 | $1.97 \times 10^9$<br>[$2.25 \times 10^7$,<br>$8.76 \times 10^{10}$] | $6.98 \times 10^9$<br>[$4.15 \times 10^7$,<br>$1.21 \times 10^{12}$] | $1.38 \times 10^9$<br>[$1.36 \times 10^7$,<br>$4.62 \times 10^{10}$] | 0.001 | 0.0 |
| Tumor Image Size | $4.60 \times 10^4$<br>[$1.19 \times 10^4$,<br>$1.30 \times 10^5$] | $4.51 \times 10^4$<br>[$1.22 \times 10^4$,<br>$1.63 \times 10^5$] | $4.60 \times 10^4$<br>[$1.19 \times 10^4$,<br>$1.24 \times 10^5$] | 0.364 | 0.0 |
| Age | 68.36 (9.78) | 69.27 (8.79) | 68.05 (10.09) | 0.206 | 3.9 |
| Stage |  |  |  | <0.001 | 0.2 |
| Stage I | 180 (32.3) | 87 (63.0) | 93 (22.1) |  |  |
| Stage II | 66 (11.8) | 26 (18.8) | 40 (9.5) |  |  |
| Stage IIIa | 131 (23.5) | 20 (14.5) | 111 (26.4) |  |  |
| Stage IIIb | 177 (31.7) | 1 (0.7) | 176 (41.9) |  |  |
| Stage IV | 4 (0.7) | 4 (2.9) | 0 (0.0) |  |  |
| Sex (% Female) | 166 (29.7) | 35 (25.4) | 131 (31.1) | 0.239 | 0.0 |
| Patient Followup | 1975.00 [1440.50,<br>2984.50] | 1834.00 [1260.00,<br>2153.00] | 3372.50 [1765.00,<br>3634.50] | <0.001 | 0.0 |

The NSCLC Radiomics dataset had a greater proportion of patients who died during its study. The mean follow up time for the censured patients (alive at the end of the study) was higher in the Radiomics cohort as well; though both feature relatively long median follow up times of at least 5 years. There was no significant difference in age between the Radiomics cohort (68.05 ± 10.09) and the Radiogenomics cohort (69.27 ± 8.79, *t* = 1.27, p × 0.206). Stage had a different distribution between the two cohorts (Stage I tumors 22.1% vs 63.0%, *χ*^2^ = 135, p < .001). There were no significant differences in sex proportion between the two data sets (*χ*^2^ = 1.38, p × 0.239). Both datasets contained a higher percentage of male patients. Moment 1 of the 0D feature curve (*Z* = 28592, p × 0.782) and tumor image size (*Z* = 30544, p × 0.36) were not significantly different between the two data sets. Both of these variables were significant in predicting survival in the univariate Cox hazard model.

We first performed a discrete analysis comparing survival quartiles by their moments of distribution of the 0D feature curves. This was performed primarily for intuition showing what the median topological feature curves looked like between survival groups. **Supplemental Figures 1, 2** and **Supplemental Table 2** show and quantify this difference.

The discrete analysis is not as rigorous as a Cox model, which would measure the impact of the moments of the 0D topological feature curve on survival outcomes. We verified that the assumptions of proportional hazards were met through visual appraisal of the Schoenfeld residuals plot, which is shown in **Supplemental Figure 4. Table 2** shows the results of our Cox proportional hazard model.

**Table 2.** Both univariate and multivariate Cox hazard models show moment 1 of the 0D feature curve is associated with poorer survival. Data is shown as hazard ratio (95% confidence interval). Bolded predictor variables were significant in both the univariate and multivariate models. Italicized predictor variables were only significant in the univariate model. The moments of distribution and tumor image size were rescaled to lie between 0 and 50 units. Linear scaling does not alter HR significance but does change the magnitude of the HR and CI to a more interpretable value. Patients with stage 0 tumors were removed from this model as the Stage 0 vs stage I variable did not converge in the model (**Supplemental Table 3**).

| Cox Proportional Hazard Model |  |  |  |  |
| --- | --- | --- | --- | --- |
|  | Univariate Model HR | p-value | Multivariate Model HR | p-value |
| **Age** | 1.018 (1.007-1.029) | 0.0019 | 1.025 (1.013-1.037) | $4.0 \times 10^{-5}$ |
| Male vs Female | 1.229 (0.994-1.52) | 0.057 | 1.128 (0.898-1.417) | 0.30 |
| **Scaled Moment 1** | 1.019 (1.005-1.033) | 0.0082 | 1.118 (1.026-1.218) | 0.011 |
| Scaled Moment 2 | 1.004 (0.982-1.026) | 0.74 | 0.766 (0.53-1.106) | 0.16 |
| Scaled Moment 3 | 0.992 (0.94-1.047) | 0.77 | 1.282 (0.412-3.991) | 0.67 |
| Scaled Moment 4 | 0.985 (0.893-1.088) | 0.77 | 0.995 (0.369-2.681) | 0.99 |
| *Scaled Tumor Image Size* | 1.015 (1.003-1.026) | 0.01 | 1.014 (0.983-1.046) | 0.38 |
| Stage II vs I | 1.249 (0.884-1.765) | 0.21 | 1.028 (0.71-1.489) | 0.88 |
| **Stage IIIa vs I** | 1.695 (1.304-2.203) | $8.0 \times 10^{-5}$ | 1.742 (1.317-2.305) | 0.0001 |
| **Stage IIIb vs I** | 1.626 (1.272-2.077) | 0.0001 | 1.483 (1.127-1.951) | 0.0049 |
| Stage IV vs I | 1.279 (0.406-4.028) | 0.67 | 1.825 (0.572-5.818) | 0.31 |

Increasing age had a significant effect on survival (multivariate HR: 1.025; 95% CI: 1.013-1.037; *z* = 4.109; p < 0.001). Sex did not have a significant effect on survival (multivariate HR: 1.128; 95% CI: 0.898-1.417; *z* = 1.037; p × 0.30). Increasing stage also had a significant effect on survival except for stage IV, which is likely due to small sample size (n × 4). Moment 1 of the 0D topological feature curves had a significant effect on survival (multivariate HR: 1.118; 95% CI: 1.026-1.218; *z* = 2.547; p × 0.011). Moment 1 of the 0D topological feature curve significantly predicted survival even after controlling for tumor image size. Tumor image size only had a significant effect on survival in the univariate model (univariate HR: 1.015; 95% CI: 1.003-1.026; *z* × 2.565; p × 0.01). **Figure 2** shows a forest plot of the Cox model. The nonsignificant Stage IV vs Stage I predictor variable was removed as its wide confidence interval interfered with data visualization.

**Figure 2.**
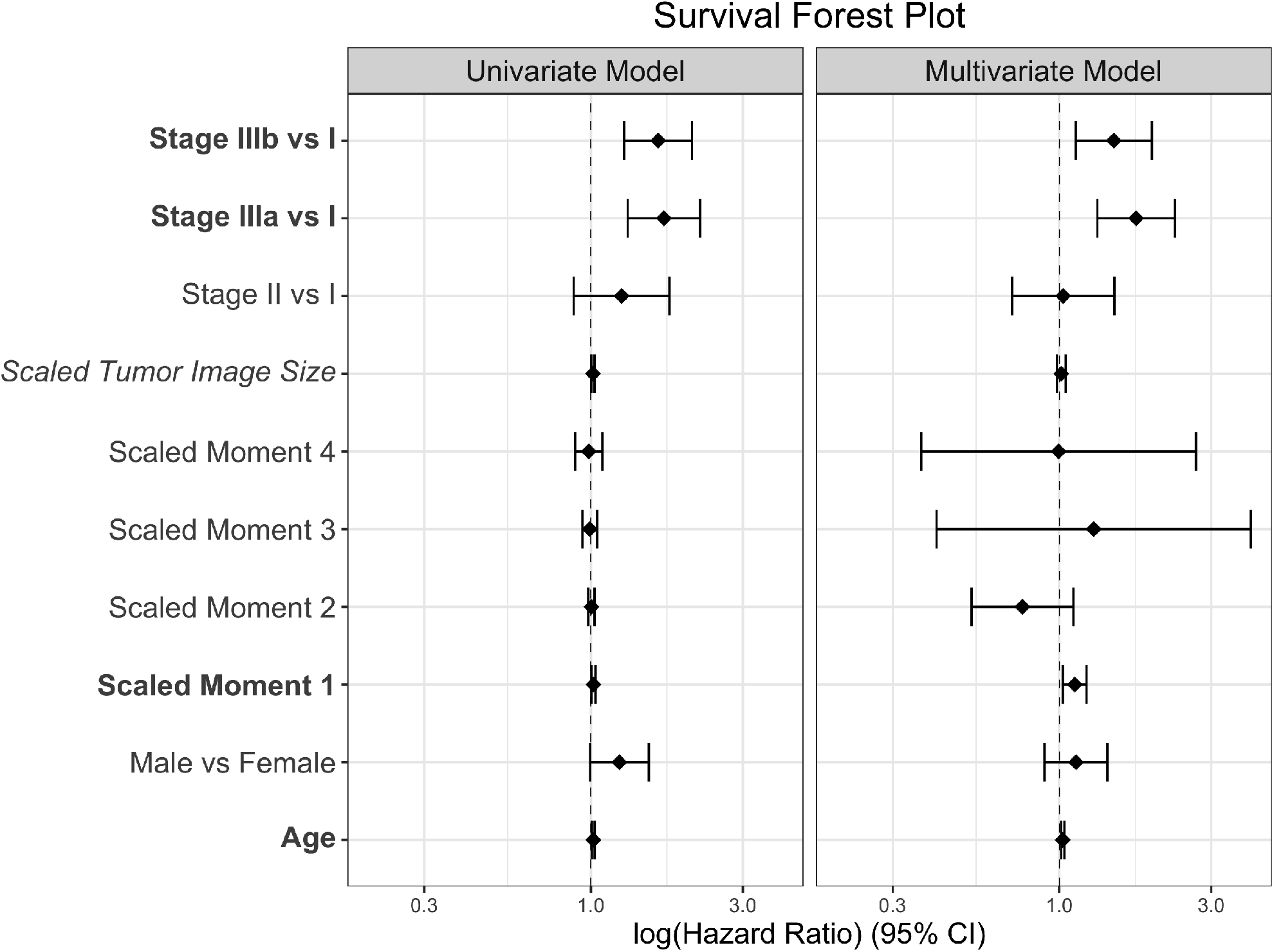
Survival forest plot shows moment 1 predicts poorer survival even after controlling for known tumor prognosticators such as stage. The horizontal axis is log transformed for better data visualization. Span of each data point represents the 95% CI. The Stage IV vs Stage I data point had a very wide CI, so it was removed from this plot for data visualization. The Stage IV vs Stage I Hazard Ratio is shown in **Table 2**. Bolded predictor variables are significant in both the univariate and multivariate models. Italicized predictor variables are only significant in the univariate model.

We used Kaplan-Meier curves (**Figure 3**) to visually justify our survival results. Patients were divided into four groups based on scaled moment 1 quartiles. The median survival is depicted by the colored vertical lines and stated with 95% CI in the legend. The four groups had significantly different survival distributions by the log-rank test (*χ*^2^ = 19.7, p < 0.001). The median survival of the first quartile (1357 days; 95% CI: 1028-1661) was significantly different from the median survival of the fourth quartile (429 days; 95% CI: 326-601; *χ*^2^ = 13.4; p × 0.0015). The median survival of the second quartile (944 days; 95% CI: 672-1490) was also significantly different from the median survival of the fourth quartile (*χ*^2^ × 12.5; p × 0.0024).

**Figure 3.**
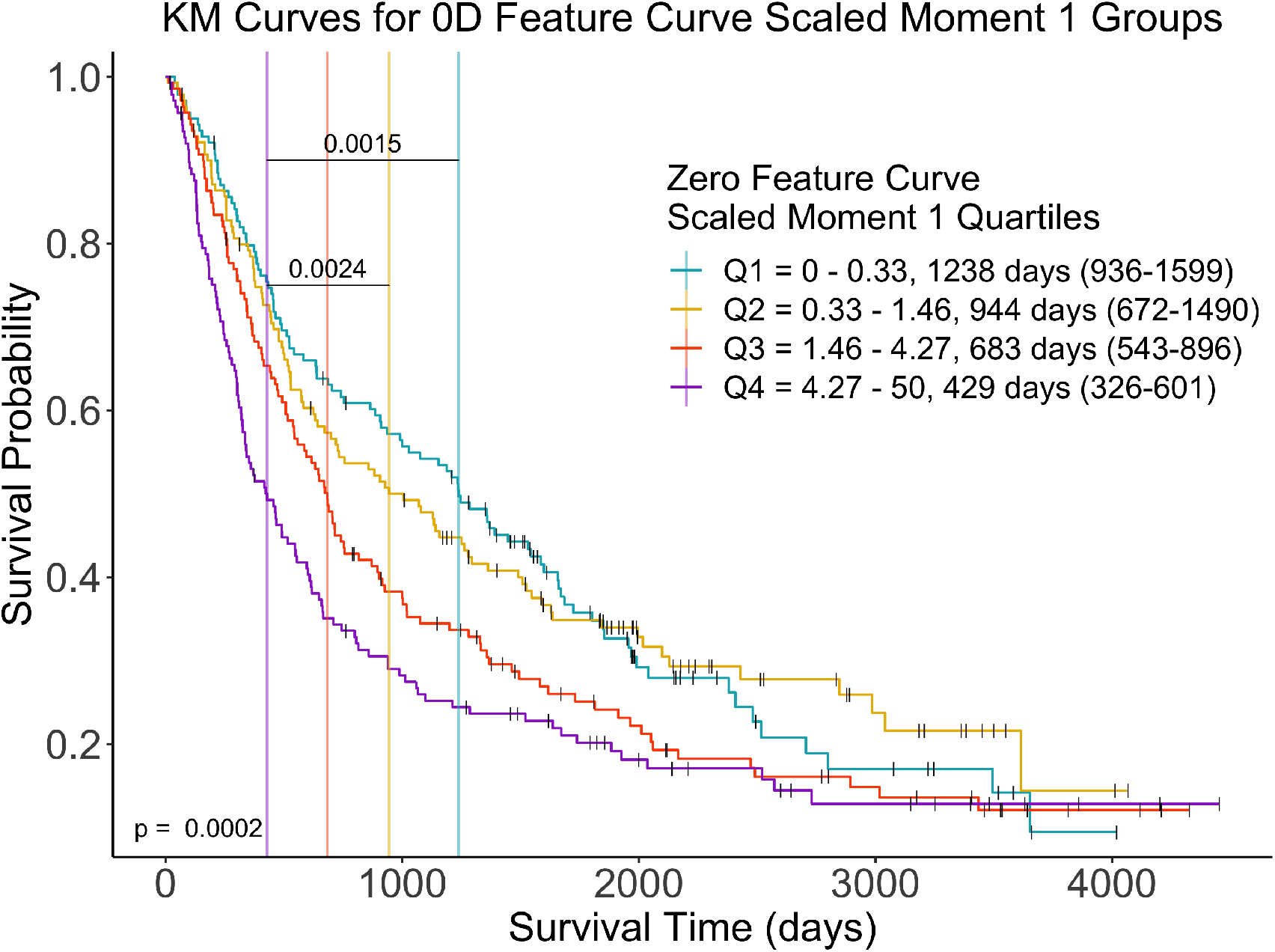
Patients with larger moment 1 values of the 0D feature curve had poorer survival. Patients were divided into quartiles based on the moment 1 value of the 0D topological feature curve. Q1 through Q4 represent Quartile 1 through Quartile 4. The exact quartile values and median survival of each quartile is shown in the figure legend. On each survival curve, the black vertical line represents a right censored event or the survival time of a patient who was still alive at last follow up. Vertical colored lines represent median survival. The result of the log-rank comparing all survival curves are shown in the bottom left. Post hoc log-rank analysis with Bonferroni correction was performed on survival curve pairs. The significant results are shown as horizontal lines connecting the median survival of the groups with significant differences.

## 4 Discussion

As the “-omics” sciences continue to become more present in the clinic, we need stronger analysis techniques that make meaning out of the massive amount of data. Compared to the other “-omics,” radiomics may be the most useful from a clinical perspective. Obtaining repeated tumor CT and MRI scans is easier for a hospital and less invasive for a patient compared to repeated tumor sequencing (genomics) or mass spectrometry (proteomics). Therefore, it would be quite valuable if persistent homology could uncover additional meaning from imaging data. We show that even after controlling for tumor stage, age, sex, and image size, the first moment of the 0D topological feature curve is a significant predictor of survival. This effect was significant across two independent data sets indicating a resilience of our methodology to batch effect. We considered tumor image size as a confounding possibility since the number of topological features would be expected to increase with image size. However, the effect of moment 1 of the 0D feature curve on survival remained significant in the multivariate model, and about 30% of the variance in moment 1 of the 0D topological feature curve was not explainable by tumor image size alone (**Supplemental Figure 5**).

It is hard to describe biological correlations of the 0D feature curve at this point. Images with a greater number of 0D features would have a more heterogenous distribution of gray scale values compared to images with a fewer number of 0D features. We conjecture that increased 0D features correlate with a tumor that is heterogeneous in tissue composition since different tissue types results in different gray scale values in a CT scan. In addition, a patchy disconnected tumor may also be reflective of an increased number of 0D features as disconnected foci of gray scale values would result in more features even if the whole tumor volume is the same compared to a well localized tumor.

Despite these promising results, there remain limitations in our study. Though the pipeline worked on a combined set of tumor scans from two separate research studies, the imaging parameters across the two sets were similar. It remains unknown how sensitive this technique is to imaging parameters such as CT machine model, which has been shown to affect calculation of standard radiomic features^16^. While we controlled for some important clinical variables as potential confounders, there are other relevant clinical variables such as EGFR gene status we were unable to test due to lack of data. We could only control for relevant clinical variables common to both data sets. In addition, while we know the analyzed scans were for planning radiotherapy or surgery, all of the patients’ prior clinical history is a black box, which is a common limitation of public databases^17,18^.

We only explored 0D topological feature curves in our analysis. We had found similar trends using other topological features curves in predicting survival. However, adding the moments of distribution of the other three feature curves would add up to 12 additional variables to consider, which we do not believe our study is adequately powered to assess. We chose to limit our focus to 0D features as they had the strongest association with survival.

Most of the aforementioned issues can be resolved by extending our pipeline to study additional CT scans from NSCLC patients and additional clinical covariates across different databases. This would allow us to control for other clinical confounders and provide greater statistical power to assess moments of distribution of higher dimensional topological feature curves.

After initial therapy, patients with lung cancer obtain follow-up CT scans to survey tumor recurrence. Local and regional failure occur respectively when the primary tumor site and nearby lesions regrow after therapy. As many of the curative intent therapies include primary-tumor directed therapy (radiation or surgery), it would be worthwhile in the future to also consider local control or local progression free survival in addition to overall survival. Long term, we envision predictive modeling that incorporates relevant clinical and radiomics variables to personalize anticancer therapy for each patient. The topological feature curve summary statistics may be appropriate variables to include in such models. Since our variable only requires a CT scan and appropriate segmentation, this tool would provide universal benefit to all healthcare practice types.

## 5 Methods

The goal of this pipeline is to determine whether calculated variables describing the topology of a tumor are associated with survival in a Cox Proportional Hazards model. Four topological variables representing 0D features or connectedness of a tumor’s CT scan were used in this Cox model. Derivation of these variables are further described in section 2

All images and segmentation objects were obtained from The Cancer Imaging Archive^13^. Within TCIA, the NSCLC-Radiomics (n × 422) and NSCLC Radiogenomics (n × 211) cohorts provided CT scans and clinical data^14,15,17,18^. Of the 633 total scans, 565 also had segmentation data, which was necessary for our pipeline. **Figure 4** shows the exclusion and inclusion criteria for each analysis.

**Figure 4.**
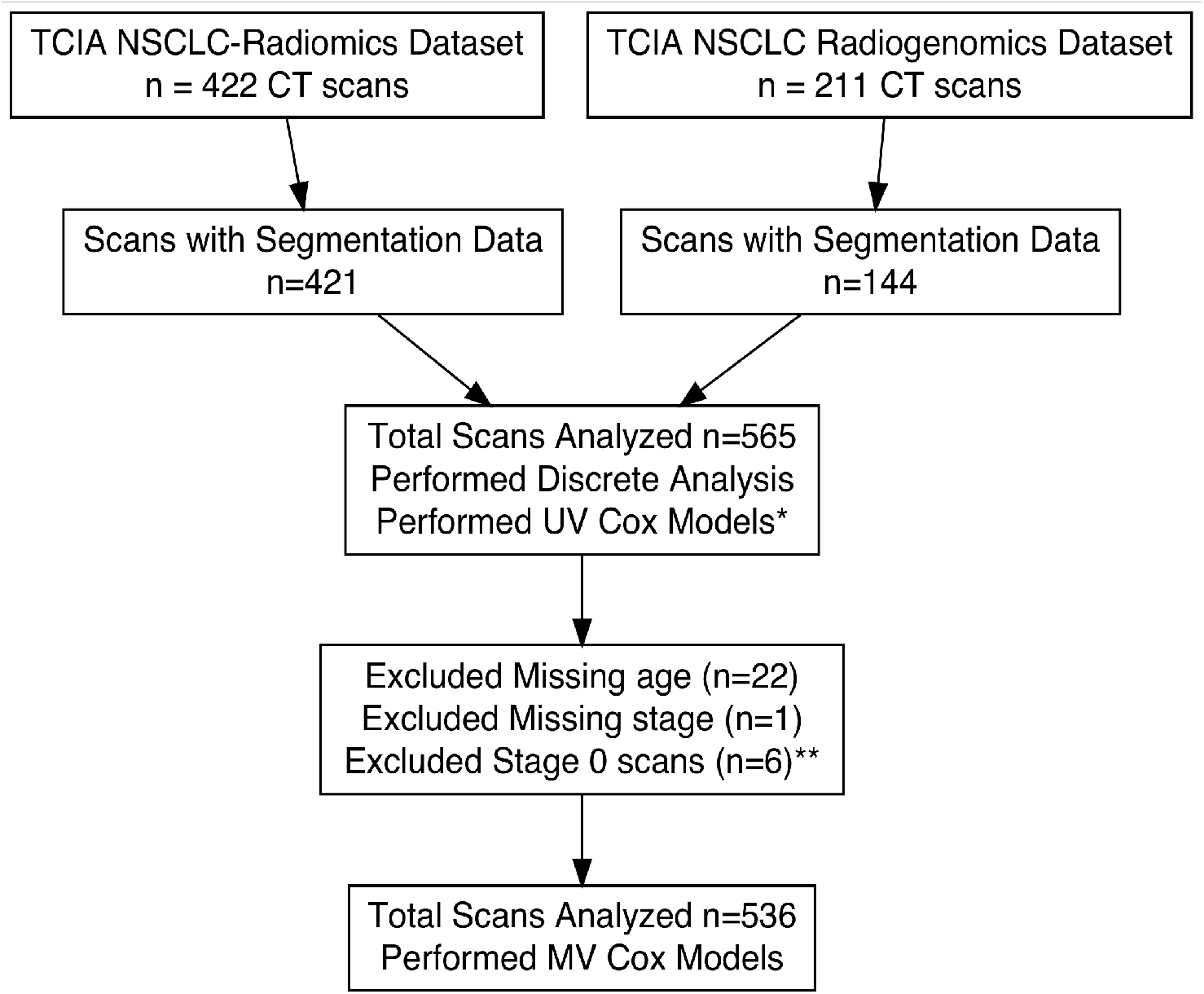
Flowchart of tumor scan inclusion and exclusion. 565 tumors had both segmentation data and CT scans allowing for computation of cubical complexes in our pipeline. *The Univariate Cox models excluded patients with missing variables. **The multivariate Cox model excluded 6 additional patients who were stage 0 since the models did not converge (**Supplemental Table 3**.

**Figure 5** shows an overview of the publicly available data pipeline using R (v3.6.1) and Python (v3.7.6) ^19–21^. Within R, a cubical region that encapsulated the primary tumor was delineated using the segmentation file coordinates of the gross tumor volume, which had been manually delineated by radiation oncologists from the original datasets. This data object was exported to Python to compute cubical complex persistence homology (functionality in R not present at time of analysis). Within R, the raw persistence homology from Python was transformed into topological feature curves, which plot the topological features against each Hounsfield filtration value. The first four moments of the 0D topological feature curve were used as predictor variables alongside clinical covariates in our survival analysis.

**Figure 5.**
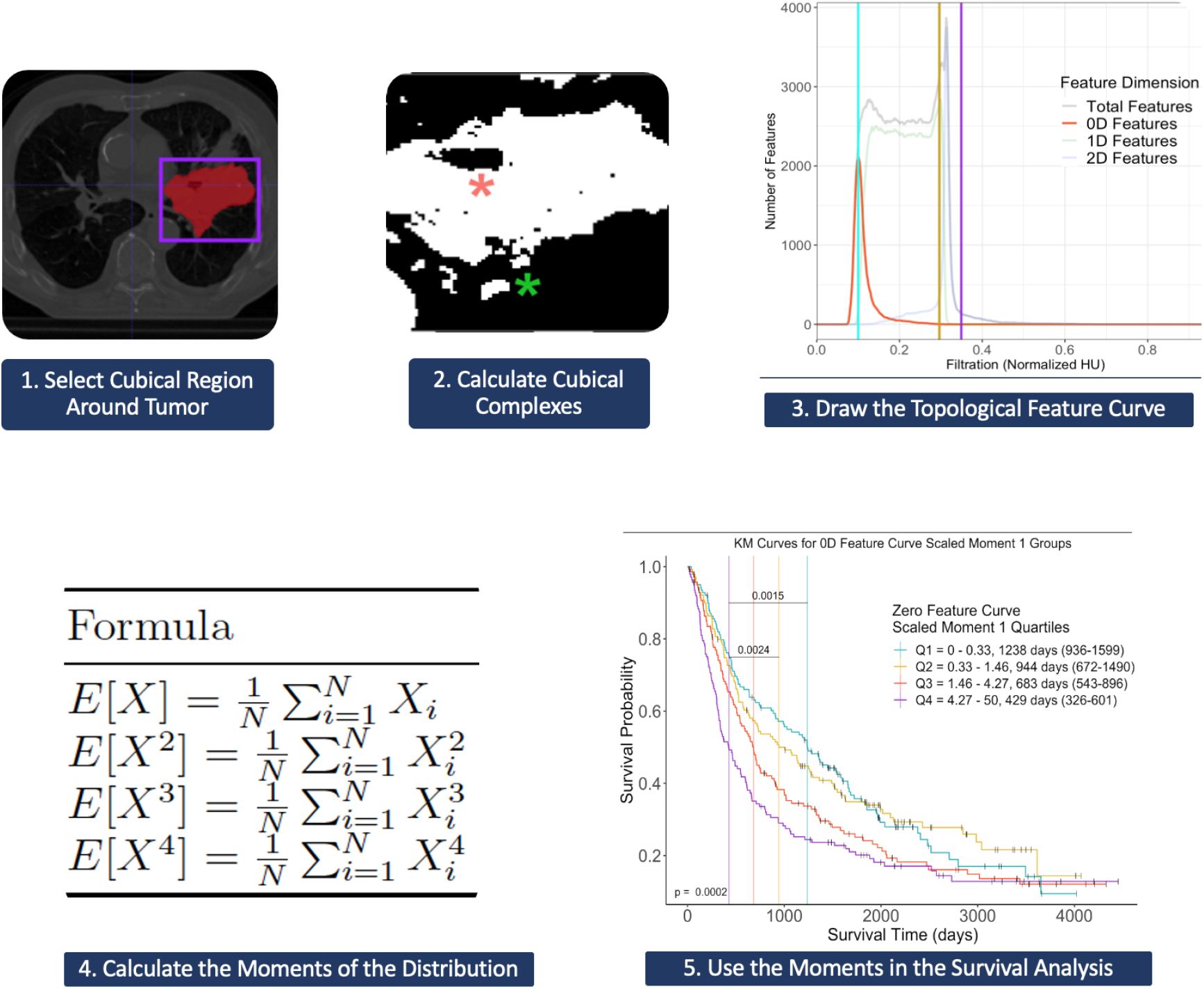
Overview of data pipeline. **(1)** Using the tumor segmentation data’s coordinates, we computed the persistent homology. **(2)** Persistent homology was computed using cubical complexes and **(3)** transformed into a feature curve as described in **Figure 1. (4)** The raw moments of the 0D feature curve were calculated and **(5)** used as predictor variables in our survival analysis (KM Curves and Cox Proportional Hazards). The pink and green asterisks in the second panel point to an example of a 0D and 1D feature in the filtered image.

For the survival analysis, univariate and multivariate Cox proportional hazard modeling was performed for the moments, tumor image size, cancer stage, age, and sex. These were all the clinical variables common to both the Radiomics and Radiogenomics datasets. Increasing the number pixels of an image likely increases the topological feature count without necessarily changing the overall topological structure of the tumor image. For this reason, we included tumor image size (pixels per slice x number of slices) as a covariate in the multivariable Cox Hazard model to control for potential confounding with our 0D topological feature curve. All survival times were directly provided in the NSCLC-Radiomics data set. This survival time was not directly provided in the NSCLC-Radiogenomics data set, but the date of imaging and date of death or last follow-up were provided. Survival times in the NSCLC-Radiogenomics were calculated by subtracting the date of death/last follow-up from date of imaging. The stage 0 predictor variable did not converge since none of the 6 patients with stage 0 cancer died during the study, so these patients were excluded (**Supplemental Table 3**).

Log-rank test was used to compare all four moment 1 quartiles in the Kaplan Meier curves, and a post hoc log-rank test was used to make pairwise comparisons. Unpaired t-test statistics (for normally distributed data) and Wilcoxon rank sum tests (for non-normally distributed data) were used to descriptively compare patient characteristics between the Radiomics and Radiogenomics data sets. Chi-squared test was used to compare categorical variables.

An open-source, reproducible pipeline is available on GitHub with detailed instructions. All packages and libraries used were open source. In R, we used `oro.dicom` v0.5.3, `oro.nifti` v0.10.3, and `RNifti` v1.1.0 to read and process the CT DICOM scans and NIfTI segmentation objects^22–24^; `dplyr` v1.0.0, `plyr` v1.8.6, and `reshape` v0.8.8, for data wrangling^25–27^; `reticulate` v1.1.6 to convert R data structures to compatible .npy files for the Python scripts^28^; `survival` v3.2-3, `rstatix` v0.6.0, and `survminer` v0.4.7 for survival analysis and statistics^29–31^; `tableone` v0.11.2 to generate table one data^32^; and `ggplot2` v3.3.2, `TDAstats` v0.4.1, `grid` v3.6.3, `png` v0.1-7, `ggplotify` v0.0.5, `ggpubr` v0.4.0, `gt` v0.2.2, `paletteer` v1.2.0, `DiagrammeR` v1.0.6.1, `DiagrammeRsvg` v0.1, `ggfortify` v0.4.10, `rsvg` v2.1, and `gridExtra` v2.3 for data visualization and export^33–45^.

In Python, we used the built in `glob` and `pathlib` libraries to recursively read in the file objects produced from the R code; `numpy` v1.18.5 to create data structures that represented the processed tumor scans numpy objects from R^46^; `gudhi` v3.0.0 to compute the persistent homology using cubical complexes^47^; and csv v1.0 library to output the computed persistent homology as csv files to be processed again in R.

We used the dcmqi open source Bash library^48^ to convert segmentation files into a more compatible NIfTI file format. We used ITK-SNAP v3.8.0 as the DICOM viewer to visually ensure our cubical segmentation properly highlighted the tumor region of interest^49^.

## Data Availability

All data and code to perform the analyses can be found at https://github.com/eashwarsoma/TDA-Lung-Phom-Reproducible.

## Supplemental Information

**Supplemental Figure 1.**
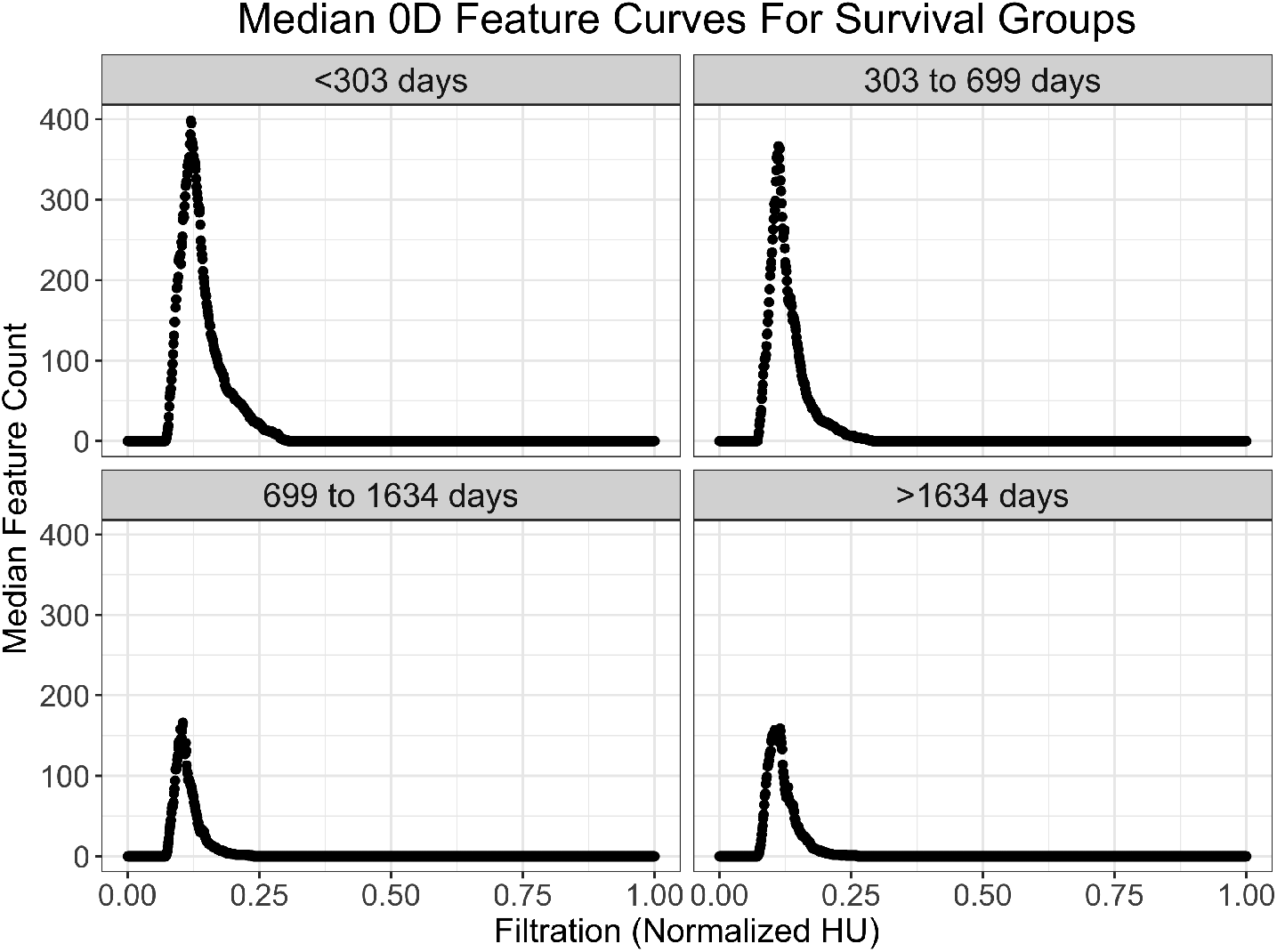
Median 0D feature curves of survival quartiles. Median 0D feature curves are larger for patients in poorer survival quartiles. Each survival group had approximately one quarter of the combined study cohorts (n × 145, n × 138, n × 141, n × 141, from worst to best survival). Within each survival group, the median 0D feature count value for each Hounsfield units was collected into one vector and plotted. The Hounsfield units were normalized to 0 to 1.

**Table 1.** Formulas to calculate the moments of the 0D feature curve. *X* refers to the vector of values represented by the vertical axis in our 0D topological feature curves. *N* refers to the number of values in that vector. *E* refers to the expected value of the distribution.

| Moment | Formula |
| --- | --- |
| Moment 1 | $E[X] = \frac{1}{N} \sum_{i=1}^N X_i$ |
| Moment 2 | $E[X^2] = \frac{1}{N} \sum_{i=1}^N X_i^2$ |
| Moment 3 | $E[X^3] = \frac{1}{N} \sum_{i=1}^N X_i^3$ |
| Moment 4 | $E[X^4] = \frac{1}{N} \sum_{i=1}^N X_i^4$ |

**Supplemental Figure 2.**
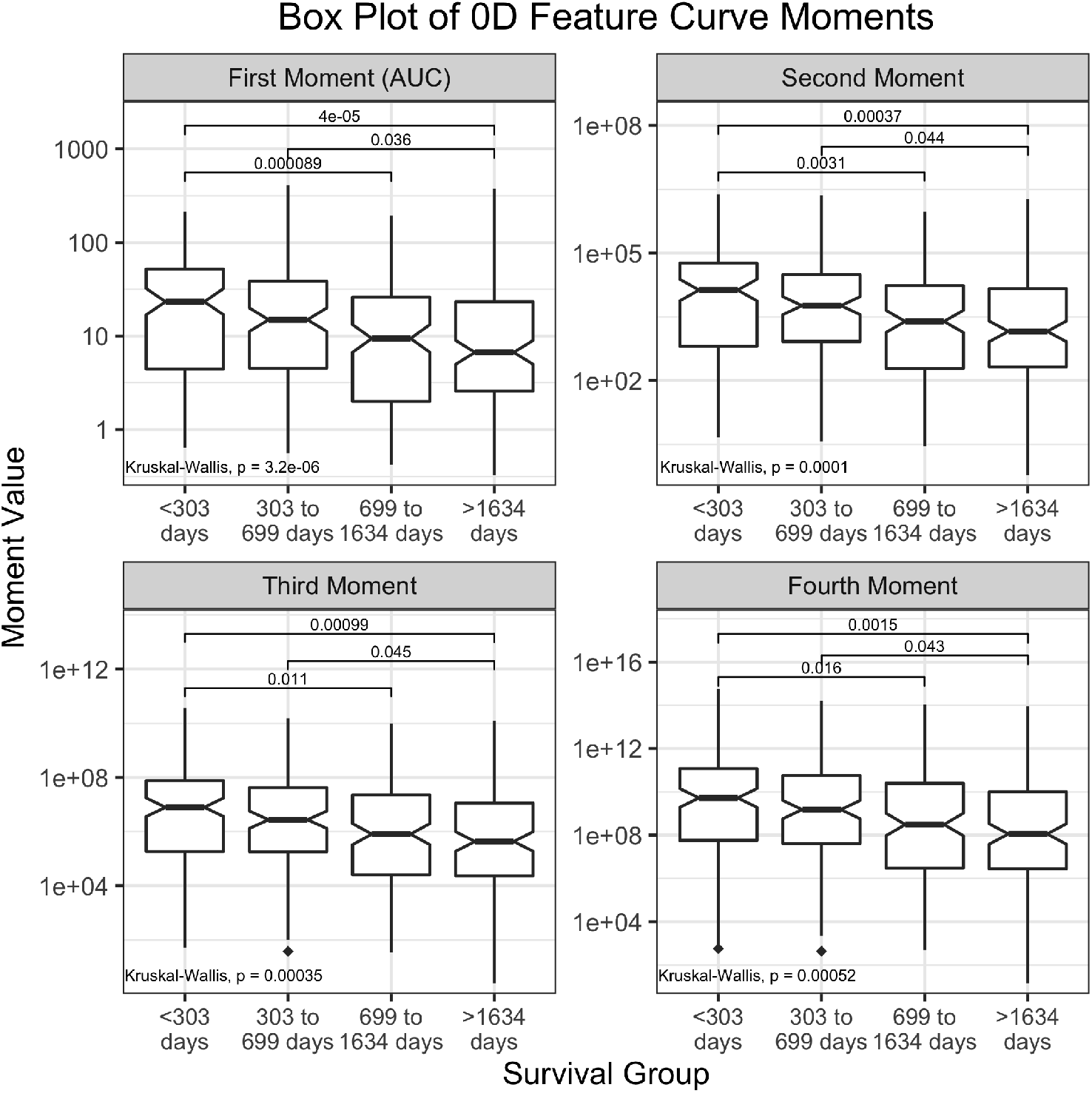
Box plots of 0D Feature Curve Moments. 0D feature curve moments are significantly higher for poorer survival quartiles as shown by box plots. The moments of distribution were analyzed for each survival group’s 0D feature curves. For data visualization, the log of the moment values are presented in the vertical axis. Each facet’s vertical axis has a different scale. Points on the plot represent outliers. Notches in the box represent a 95% confidence interval around the median. Kruskal-Wallis H Test was performed to compare median value for each moment among the survival groups, and post hoc Dunn’s test with Bonferroni correction was performed to compare pairs of survival groups. Exact median moment values and interquartile ranges are provided in **Supplemental Figure 3**. Exact Kruskal-Wallis and post hoc Dunn’s test comparisons are found in **Supplemental Table 2**.

**Table 2.**
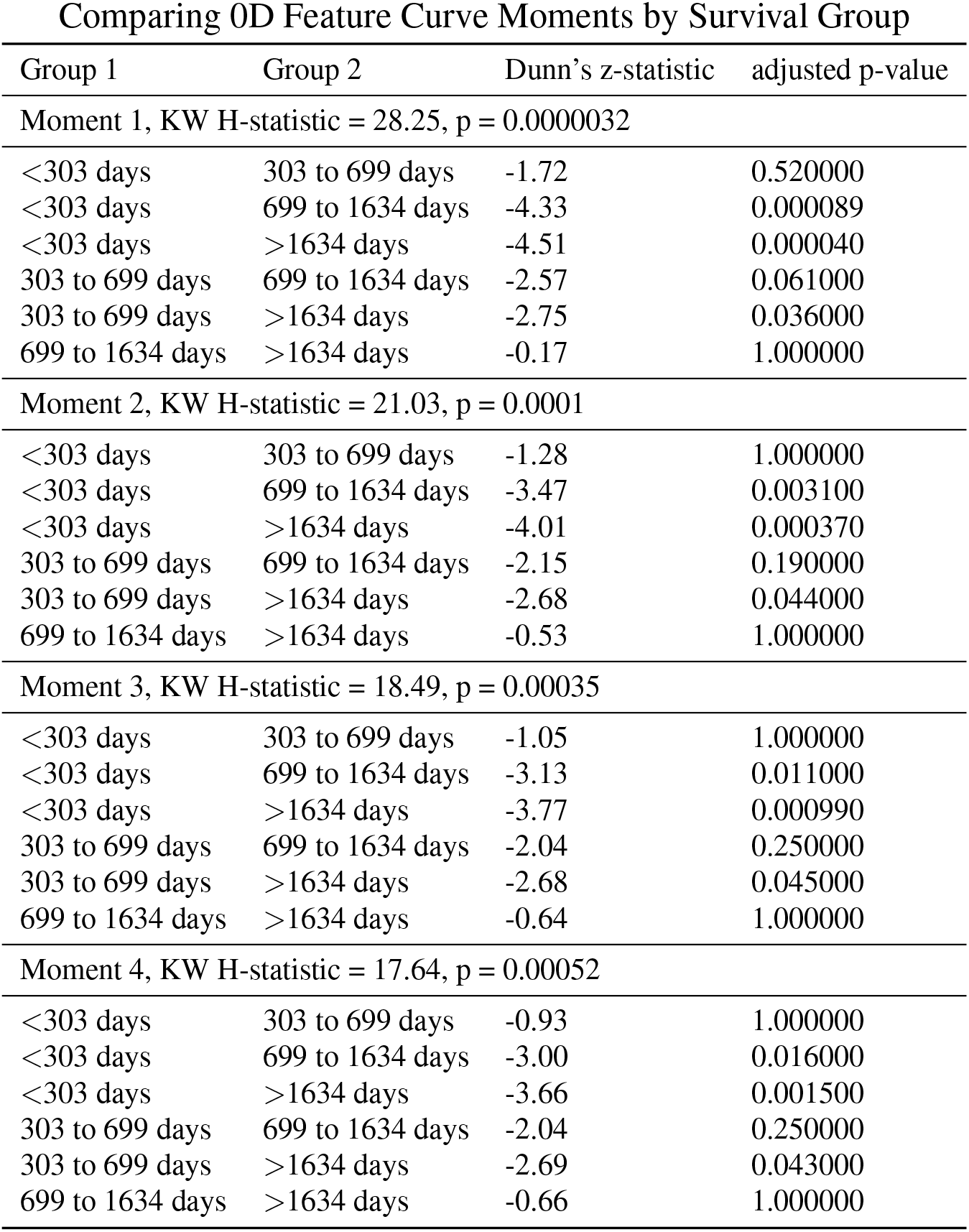
Statistical comparisons between each survival group for each moment. Kruskal-Wallis H test was used to compare survival groups within each moment variable. Post hoc Dunn’s test was performed to make pairwise comparison between survival groups. Bonferroni adjusted p-values are shown.

**Table 3.** Cox table with stage 0 patients not excluded. Stage 0 patients were excluded in the original analysis as the stage 0 vs stage I variable did not converge since no stage 0 patients died. Including stage 0 patients does not impact any of the conclusions of the original analysis.

|  | Univariate Model HR | p-value | Multivariate Model HR | p-value |
| --- | --- | --- | --- | --- |
| Age | 1.018 (1.007-1.029) | 0.0018 | 1.025 (1.013-1.037) | $4.0 \times 10^{-5}$ |
| Male vs Female | 1.212 (0.98-1.499) | 0.0770 | 1.128 (0.898-1.417) | 0.3000 |
| Scaled Moment 1 | 1.019 (1.006-1.033) | 0.0054 | 1.118 (1.026-1.218) | 0.0110 |
| Scaled Moment 2 | 1.005 (0.983-1.027) | 0.6900 | 0.766 (0.53-1.106) | 0.1600 |
| Scaled Moment 3 | 0.994 (0.941-1.049) | 0.8100 | 1.282 (0.412-3.991) | 0.6700 |
| Scaled Moment 4 | 0.988 (0.895-1.09) | 0.8100 | 0.995 (0.369-2.681) | 0.9900 |
| Scaled Tumor Image Size | 1.015 (1.003-1.026) | 0.0100 | 1.014 (0.983-1.046) | 0.3800 |
| Stage 0 vs I | 0 (0-Inf) | 0.9900 | 0 (0-Inf) | 0.9900 |
| Stage II vs I | 1.249 (0.884-1.765) | 0.2100 | 1.028 (0.71-1.489) | 0.8800 |
| Stage IIIa vs I | 1.695 (1.304-2.203) | $8.0 \times 10^{-5}$ | 1.742 (1.317-2.305) | 0.0001 |
| Stage IIIb vs I | 1.626 (1.272-2.077) | 0.0001 | 1.483 (1.127-1.951) | 0.0049 |
| Stage IV vs I | 1.279 (0.406-4.028) | 0.6700 | 1.825 (0.572-5.818) | 0.3100 |

**Supplemental Figure 3.**
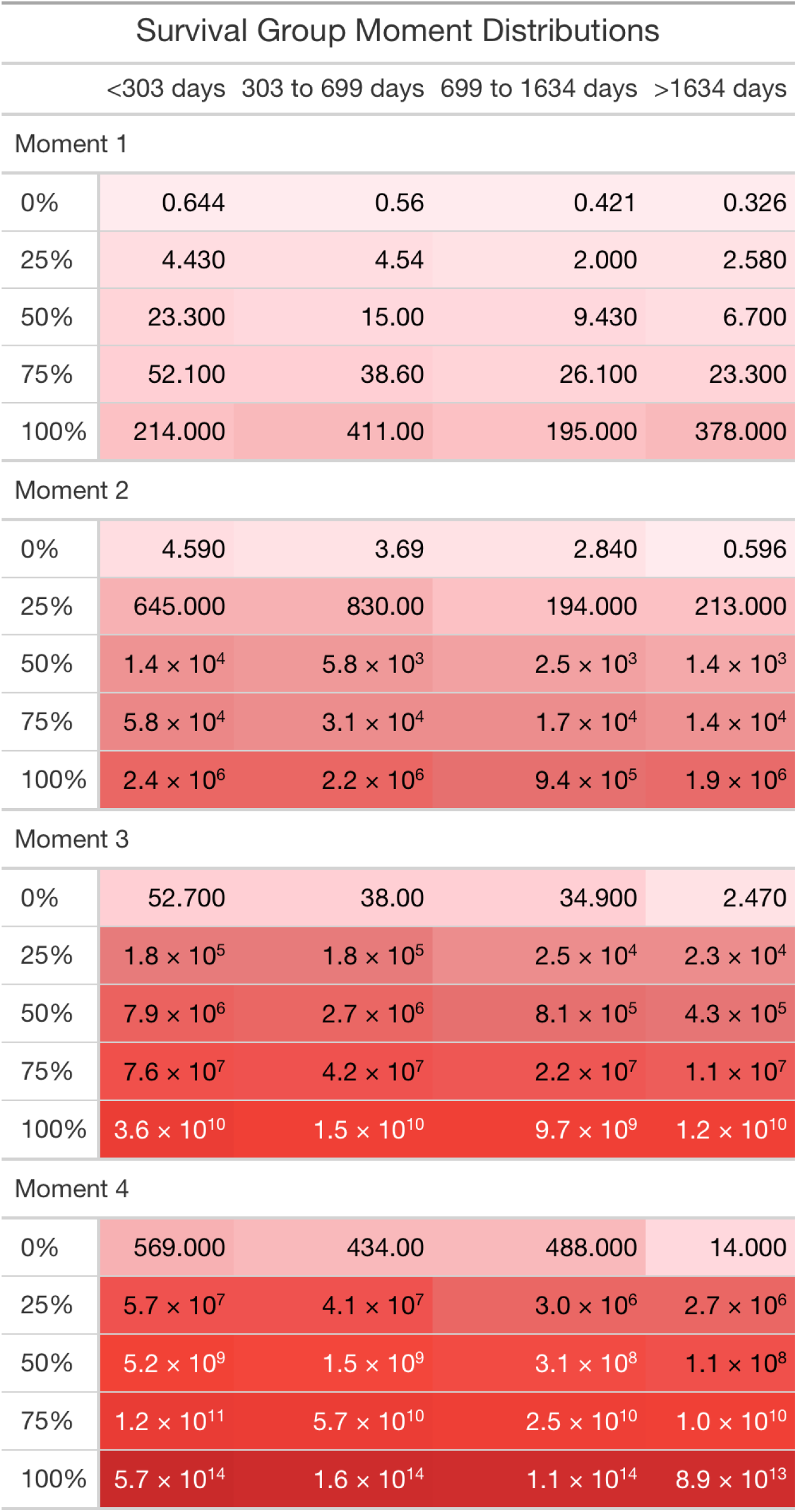
Exact raw moment values for each survival group. Groups were divided into quartiles based on survival. Each 0D feature curve was analyzed for the first four moments of distribution. The range of the moment values for each group is provided here. Color depth represents magnitude of moment value.

**Supplemental Figure 4.**
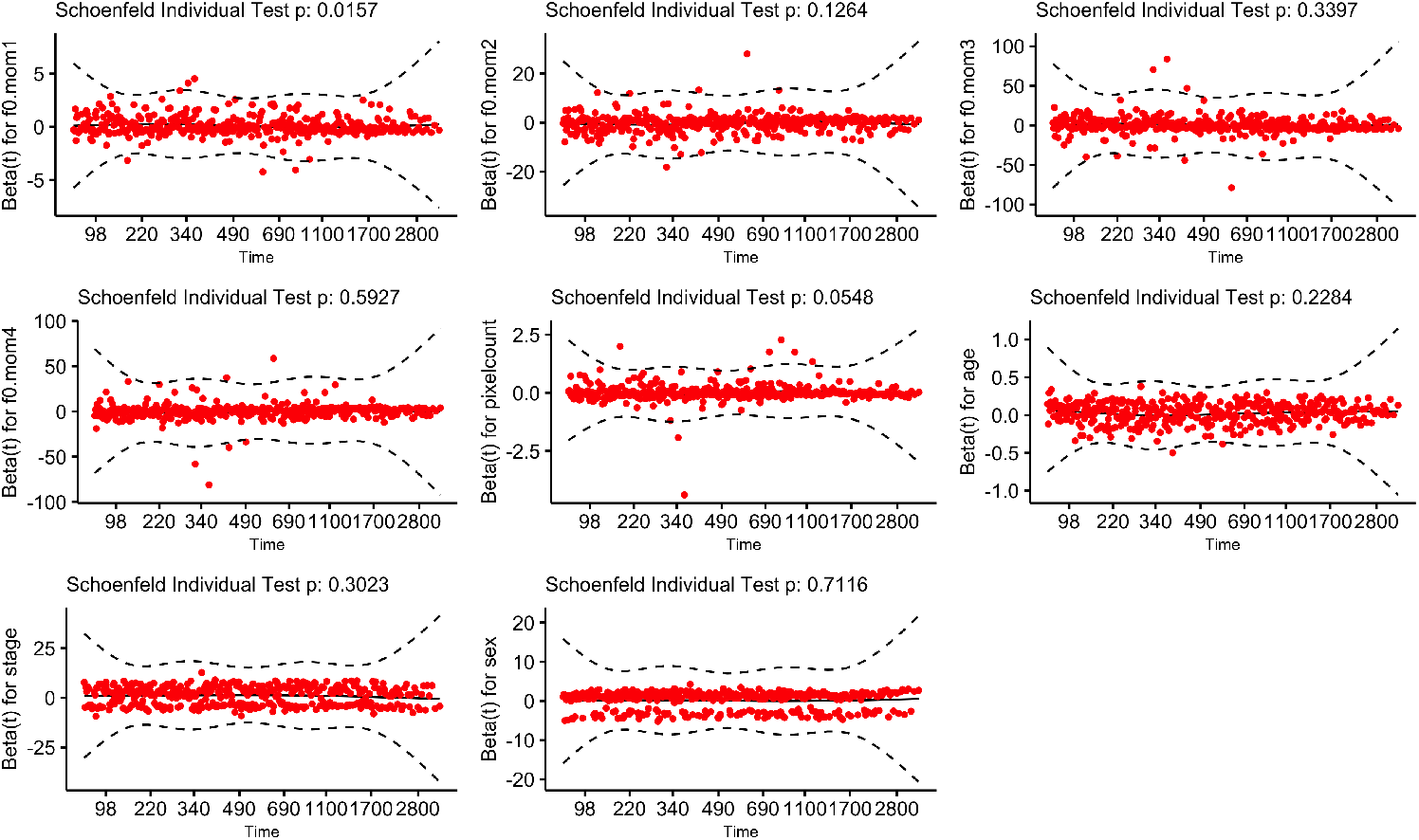
Schoenfeld residuals plot. Schoenfeld residuals were visually appraised to ensure assumptions of proportionality were met. While moment 1’s Schoenfeld residuals in the figure are significant, this is likely a chance result as visually there does not seem to be a trend in the residuals over time. The global Schoenfeld residuals were also nonsignificant

**Supplemental Figure 5.**
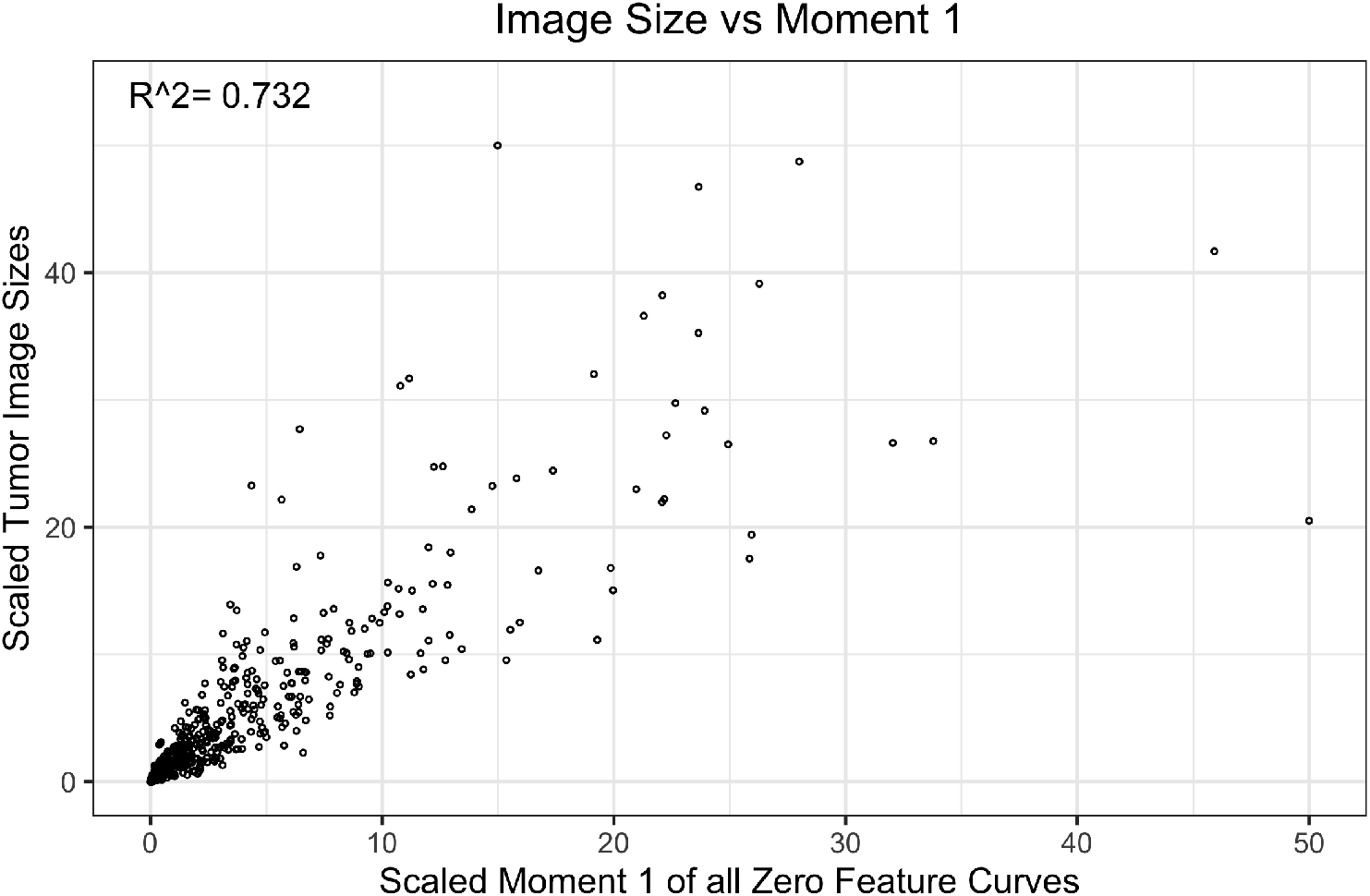
Correlation plot comparing tumor image size and moment 1 of 0D topological feature curves. Increasing the image size is expected to increase the topological features. Tumor image size was calculated by multiplying the number of pixels per CT scan slice by the number of slices. Both variables were linearly scaled to 0 to 50. We generated a correlation plot to check how much variance provided by moment 1 of the 0D feature curve was unexplained by tumor image size. About 30% of the variance in moment 1 is unexplained by tumor image size. Despite the large overlap, moment 1 remained significant even after controlling for tumor image size in our Cox model.

## Acknowledgements

We would like to thank Theory Division for their constructive feedback and valuable input in this project. Funding for this project was provided by the Case Comprehensive Cancer Center medical student summer research training grant.

## Author contributions statement

E.S. conceived the project. E.S., A.L., and R.W. designed project plan. E.S. and A.L. wrote code for the project. E.S. wrote manuscript. J.G.S. and S.O. supervised all previous steps. All authors reviewed and revised the manuscript.

